# Peri-Lead Edema in Deep Brain Stimulation: Long-Term Outcomes and Possible Etiological Correlates

**DOI:** 10.1101/2025.05.01.25326737

**Authors:** Luigi G. Remore, Giorgio Fiore, Elena Pirola, Linda Borellini, Francesca Mameli, Fabiana Ruggiero, Eleonora Zirone, Roberta Ferrucci, Filippo Cogiamanian, Enrico Mailland, Antonella M. Ampollini, Giulio A. Bertani, Stefania E. Navone, Giovanni Marfia, Ioannis U. Isaias, Marco Locatelli

**Affiliations:** Neurosurgery Unit, Fondazione IRCCS Ca’ Granda Ospedale Maggiore Policlinico, Milan, Italy; Department of Pathophysiology and Transplantation, University of Milan, Milan, Italy; Neuropathophysiology Unit, Fondazione IRCCS Ca’Granda Ospedale Maggiore Policlinico, Milan, Italy; Laboratory of Experimental Neurosurgery and Cell Therapy, Unit of Neurosurgery, Fondazione IRCCS Ca’ Granda Ospedale Maggiore Policlinico, Milan, Italy; Parkinson Institute of Milan, ASST G. Pini-CTO, Milan, Italy; University Hospital of Wuerzburg and Julius Maximilian University of Wuerzburg, Wuerzburg, Germany

## Abstract

**Background:** Deep brain stimulation (DBS) is an effective surgical procedure for the treatment of Parkinson’s disease (PD) and other movement disorders. Immediate and delayed complications after DBS surgery have been described. Peri-lead edema (PLE) is a DBS-related complication whose etiology is still unknown. Moreover, PLE frequency and long-term effects are subjects of ongoing debate.

**Objectives:** To elucidate the long-term clinical and neuropsychological effects of PLE and to find possible etiological correlates.

**Methods:** We retrospectively collected clinical and neuropsychological data from 51 PD patients before and one year after DBS. PLE visualized on FLAIR MRI sequence was manually segmented. Using appropriate statistical tests, continuous and categorical variables were compared between patients with and without PLE. A multivariate regression model was employed to analyze the contribution of clinical variables to edema volume changes.

**Results:** 68.62% of patients presented PLE at the immediate postoperative MRI. Patients with PLE were significantly older (p<0.001) and had more frequent postoperative confusion episodes (p=0.025). Furthermore, more MER (microelectrode recording) tracks (p<0.001) were used in patients with PLE. Multiple MER tracks were directly correlated with edema volume and were the only significant predictors of edema volume changes in a multivariate regression model. No differences were found in other clinical and neuropsychological variables.

**Conclusions:** PLE is a frequent post-surgical event and may cause transient postoperative confusion. It seems linked to older age and multiple MER tracks. Although it does not influence global motor and neuropsychological outcomes, PLE contributes to postoperative confusion episodes. To avoid PLE sequelae, using multiple MER tracks in older patients should be discouraged.

## Introduction

Deep brain stimulation (DBS) has been widely proven to be effective for the treatment of movement disorders, such as Parkinson’s disease (PD), essential tremor and dystonia^1^. Being employed in clinical practice for over thirty years, DBS has been demonstrated to reduce motor fluctuations and refractory tremor and to improve PD patients’ quality of life^2^. Moreover, its role in the management of different neuropsychiatric disorders^3^ has recently been investigated and DBS is currently approved to improve the symptoms related to drug-resistant obsessive-compulsive disorder^4^ and epilepsy^5,6^. Although it is usually considered a safe procedure, complications related to DBS have been reported in the literature^7^. The most common are hemorrhages, infections and hardware-related malfunctions^8^.

Peri-lead edema (PLE) is an underrecognized complication of DBS procedures^9^, with an increasing number of scientific reports raising awareness of this event in recent years^10^. PLE is defined as the presence of an edematous reaction around the DBS electrode^11^ with a reported incidence between 0.4 and 6.9%^12,13^. In this context, our group prospectively evaluated a cohort of PD patients and described that all patients displayed PLE at brain MR several months after the implantation^14^. PLE is usually asymptomatic^15,16^, although cases of hyperacute neurological deterioration presenting with focal neurological deficits and seizures were reported^17,18^ On the other hand, PLE did not appear to influence the neuropsychological profile of PD patients, at least on short-term follow-ups^13^.

Given the currently unknown etiology of PLE and its poorly understood impact on long-term outcomes, this study aims to investigate the clinical and neuropsychological associations of PLE in PD patients undergoing DBS of the subthalamic nucleus (STN) at the last follow-up available. Then, we will try to find possible correlates of PLE formation.

## Materials and methods

### Patients’ selection and peri-lead volume estimation

We retrospectively reviewed our institution’s clinical database and searched for PD patients who received bilateral STN-DBS and performed a brain MRI at least 24 hours after surgery. Fifty-one patients resulted from our search. For each patient, the following variables were collected: demographic data (sex, age at PD onset and implantation, educational level), clinical data (disease duration, L-Dopa [levodopa] dosage, LEDD [levodopa equivalent daily dose], MDS-UPDRS-III [Movement Disorder Society-Unified Parkinson’s Disease Rating Scale part III] score, the MoCA [Montreal Cognitive Assesment] total score and domain-specific sub-scores: memory, visuospatial abilities, executive functions, attention, language and orientation^19,20^). Clinical variables were reported before and one year after the implantation.

For each patient, MRI was screened for PLE, which was identified as signal hyperintensity surrounding the electrode on FLAIR MRI sequences. With the commercially available software BrainLab, PLE was manually segmented by two expert users and its volume was calculated. Segmentation results were checked by two experienced neurosurgeons. Then, patients were divided into two groups based on the presence or absence of PLE.

All PD patients included in this study were followed up by attending neurologists and neuropsychologists with many years of experience in the field of movement disorders. The surgical procedures were performed by the same attending neurosurgeons at the Neurosurgery Department of our institution.

### Surgical procedure

Our surgical technique consisted of a two-step procedure as previously described in other publications^21,22^. Briefly, the first step was conducted under local anaesthesia in the awake patient. Before surgery, a Cosman-Roberts-Wells stereotactic frame (Radionics by Integra, Plainsboro, New Jersey, USA) was placed on the patient’s head and a stereotactic CT scan was performed. A dedicated workstation (Istereotaxy by BrainLab, Kapellenstrat, Germany) was used to co-register the CT scan with the MRI performed the day before to plan STN surgical targeting. An MER (microelectrode recording) (microTargeting Electrode^TM^, FHC Inc, Bowdoin, Maine, USA) was inserted before lead implantation and the electrophysiologic activity of STN was tested. Then, the neurologist checked for possible side effects during intraoperative stimulation. In case of poor STN recording or side effects at low amplitude stimulation, a second MER track was tested. Finally, DBS leads (Model 3389 by Medtronic Inc., Minneapolis, Minnesota, USA) were implanted bilaterally. A second surgery was scheduled within seven days after the first one and performed under general anesthesia. During this surgical time, intracranial electrodes were elongated with lead extenders, which were then connected to an IPG Medtronic Activa PC implanted into a subcutaneous pouch in the right hemithorax. Patients were subsequently discharged and the stimulation was turned on three to six weeks after surgery during a second hospitalization.

### Statistical analysis

Categorical variables were reported as absolute numbers and percentages, while continuous variables were reported as mean plus standard deviation. A Shapiro-Wilk normality test was used to assess the normality across selected variables along with a close inspection of the data plotted on histograms and Q-Q graphs. A Chi-Square test was used to compare categorical variables between patients with and without PLE. MANOVA was employed to perform multivariate comparisons between the two groups of patients. In this context, the Pillai-Bartlett trace (V) multivariate test was used as a test for overall statistical significance among all variables and ANOVA univariate F-statistic as a follow-up test in case of multivariate significance. Pearson’s correlation (r) was used to examine relationships between two continuous variables and standard multivariate regression to account for the possible effect of several clinical variables on the change of a chosen variable. Confidence intervals were bootstraped by 1000 samples. For all traditional hypotheses, p values < 0.05 were considered statistically significant. Statistical analysis was performed using SPSS (version 29.0, IBM).

## Results

### Overall patients’ demographics

Fifty-one PD patients were included in this study and one hundred and two electrodes were implanted in STN bilaterally. Most patients were female (26/51, 51%). The mean PD duration before surgery and mean age at implantation were 11.53 ± 3.52 and 55.61 ± 10.07 years, respectively. The mean educational level – i.e., years spent for education purposes-was 12.5 ± 3.82. Other clinical and neuropsychological results from preoperative and one-year follow-up evaluations are reported in **Table 1**.

**Table 1:** Clinical and neuropsychological variables measured at the baseline before the surgery and at a one-year follow-up visit.

| **Variable** | **Baseline** | **One-year follow-up** |
| --- | --- | --- |
| L-Dopa<sup>1</sup> | 855.75 ± 418.11 | 419.86 ± 290 |
| LEDD<sup>2</sup> | 1150 ± 363 | 572 ± 330 |
| MDS-UPDRS-III OFF<sup>3</sup> | 40.73 ± 11.61 | 22.8 ± 8.86 |
| MoCA total<sup>4</sup> | 25.43 ± 2.54 | 25.66 ± 3 |
| MoCA Memory | 2.47 ± 1.5 | 2.94 ± 1.46 |
| MoCA Visuospatial functions | 3.31 ± 0.81 | 3.22 ± 0.8 |
| MoCA Esecutive functions | 3.55 ± 0.75 | 3.45 ± 0.85 |
| MoCA Attention | 5.51 ± 0.64 | 5.43 ± 0.8 |
| MoCA Language | 5.57 ± 0.6 | 5.43 ± 0.75 |
| MoCA Orientation | 5.88 ± 0.47 | 5.92 ± 0.27 |

1: levodopa dosage in mg. 2: levodopa equivalent daily dose before the implantation in mg/day. 3: Movement Disorder Society-Unified Parkinson's Disease Rating Scale part III. 4: Montreal Cognitive Assessment score.

After a mean postoperative time of 8.4 ± 6.37 days, the mean total PLE volume was 12.37 ± 14.85 mm^3^, while the mean total volume of hemorrhagic spots around the lead was 0.5 ± 1.75 mm^3^. There were no symptomatic hematomas requiring surgical treatment in our cohort.

### Edema groups comparison

At the first postoperative MRI, 68.62% of patients (35/51) displayed PLE (**Figure 1**). There were more females among patients with postoperative PLE (21/35, 60% VS 5/16, 31.3%; p=0.057). Postoperative confusion was more common in patients with PLE (9/35, 25.7% VS 0/16, 0%, p=0.025).

**Figure 1:**
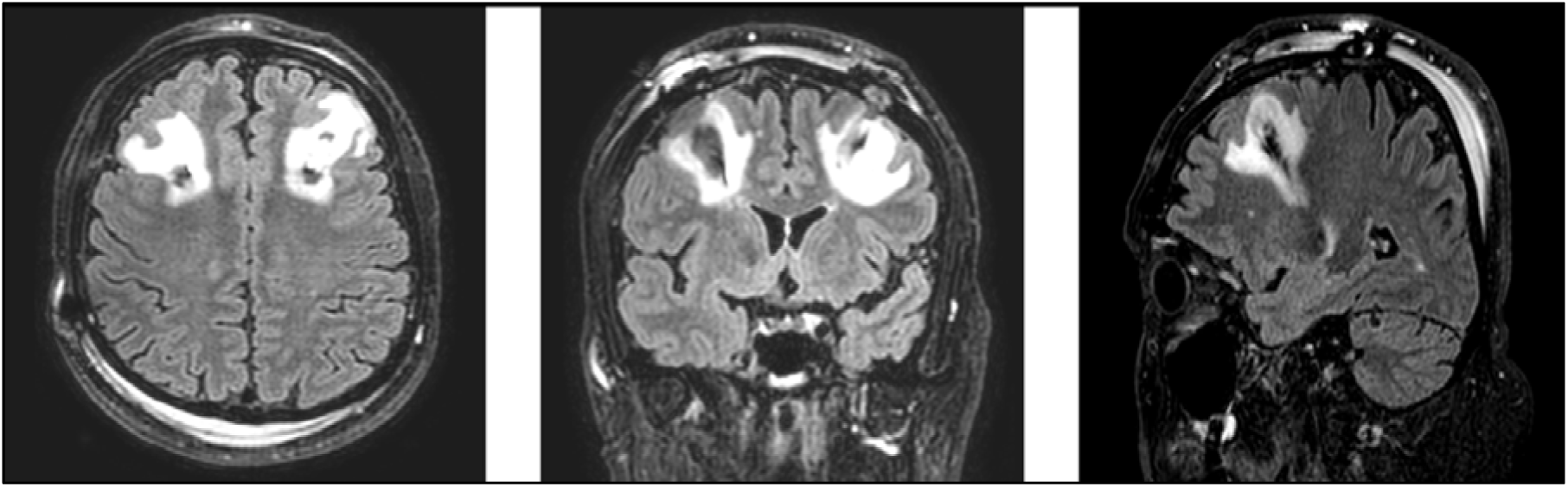
Peri-lead edema on brain MRI after STN-DBS Illustrative case of an implanted patient displaying bilateral peri-lead edema at the first post-operative MRI scan. In the FLAIR sequence, the hyperintense signal surrounding the electrode is visible from the cortical surface to the deep white matter. Brain scan projections from left to right: axial, coronal and sagittal.

When comparing clinical and neuropsychological evaluations between patients with and without edema, the multivariate test was statistically significant (V=0.83, F(24,26)=4.7, p<0.001). The follow-up univariate analysis showed that PLE patients were significantly older (p<0.001), received more MER tracks during surgery (p<0.001) and had lower dosages of parkinsonian drugs both before and after the implantation: L-Dopa pre-(p=0.002) and postoperative (p=0.037); LEDD pre-(p=0.016) and postoperative (p=0.002). No statistically significant differences were found for the other variables (**Table 2**).

**Table 2:** Comparison of the clinical and neuropsychological variables between patients with and without perielectrode edema.

| Variable | Edema | No Edema | F<sup>10</sup> | p-value |
| --- | --- | --- | --- | --- |
| Age at implantation<sup>1</sup> | 59.34 ± 7.8 | 47.44 ± 9.86 | 21.6 | <0.001<sup>*</sup> |
| Disease duration<sup>2</sup> | 11.91 ± 3.65 | 10.7 ± 3.17 | 1.34 | 0.25 |
| MER Track<sup>3</sup> | 3.31 ± 0.86 | 2 ± 0.0 | 36.4 | <0.001<sup>*</sup> |
| Hemorrhage spots volume | 0.72 ± 0.0 | 0.0 ± 0.0 | 1.92 | 0.17 |
| MRI Days<sup>4</sup> | 8.9 ± 4.9 | 7.31 ± 8.91 | 0.66 | 0.41 |
| L-Dopa<sup>5</sup> preoperative | 738 ± 343.51 | 1113.4 ± 460.5 | 10.54 | 0.002<sup>*</sup> |
| L-Dopa at one-year follow-up | 408.52 ± 43.6 | 471.7 ± 65.73 | 4.57 | 0.037<sup>*</sup> |
| LEDD<sup>6</sup> preoperative | 1056.6 ± 328.8 | 1108.1 ± 406.3 | 6.28 | 0.016<sup>*</sup> |
| LEDD at one-year follow-up | 460 ± 242 | 758.25 ± 396 | 11.02 | 0.002<sup>*</sup> |
| MDS-UPDRS-III<sup>7</sup> OFF preoperative | 40 ± 10.75 | 43 ± 13.43 | 0.08 | 0.85 |
| MDS-UPDRS-III OFF at one-year follow-up | 22 ± 8.56 | 25 ± 9.4 | 1.28 | 0.26 |
| MoCA<sup>8</sup> total preoperative | 25.5 ± 0.4 | 25.6 ± 0.6 | 0.66 | 0.42 |
| MoCA total at one-year follow-up | 26 ± 2.64 | 25.4 ± 4 | 0.21 | 0.65 |
| MoCA Memory preoperative | 2.5 ± 1.4 | 2.44 ± 1.8 | 0.01 | 0.91 |
| MoCA Memory at one-year follow-up | 2.91 ± 1.31 | 3 ± 1.8 | 0.03 | 0.84 |
| MoCA Visuospatial functions preoperative | 3.31 ± 0.83 | 3.31 ± 0.8 | 0.00 | 0.99 |
| MoCA Visuospatial functions at one-year follow-up | 3.23 ± 0.7 | 3.2 ± 0.98 | 0.03 | 0.86 |
| MoCA Esecutive functions preoperative | 3.66 ± 0.54 | 3.31 ± 1.07 | 2.34 | 0.13 |
| MoCA Esecutive functions at one-year follow-up | 3.54 ± 0.65 | 3.25 ± 1.18 | 1.3 | 0.26 |
| MoCA Attention preoperative | 5.63 ± 0.54 | 5.25 ± 0.7 | 4 | 0.05 |
| MoCA Attention at one-year follow-up | 5.5 ± 0.81 | 5.31 ± 0.8 | 0.5 | 0.5 |
| MoCA Language preoperative | 5.6 ± 0.5 | 5.5 ± 0.73 | 0.33 | 0.57 |
| MoCA Language at one-year follow-up | 5.46 ± 0.65 | 5.4 ± 0.95 | 0.13 | 0.72 |
| MoCA Orientation preoperative | 5.67 ± 0.55 | 5.94 ± 0.25 | 0.31 | 0.6 |
| MoCA Orientation at one-year follow-up | 5.9 ± 0.32 | 6 ± 0.0 | 2 | 0.16 |
| Educational Level<sup>9</sup> | 12.71 ± 3.5 | 12 ± 4.6 | 0.37 | 0.54 |

1: mean age in years at the implantation. 2: mean duration of Parkinson's disease in years until the surgery; 3: mean number of microelectrode recordings (MER) tracks used for both sides during the surgery. 4: mean number of postoperative days before the brain MRI. 5: levodopa dosage in mg. 6: levodopa equivalent daily dose before the implantation in mg/day. 7: Movement Disorder Society-Unified Parkinson's Disease Rating Scale part III. 8: Montreal Cognitive Assessment score. 9: mean years spent for education purposes. 10: F-statistics and p-value reported from follow-up univariate comparison among the two groups' clinical variables using ANOVA. \*p<0.05 was considered statistically significant.

### Predictors of edema volume

The total edema volume was directly correlated with the number of MER tracks used (r = 0.65, 95% BCa CI [0.51, 0.8], p<0.001) and inversely correlated with antiparkinsonian drugs: preoperative L-Dopa (r = -0.312, 95% BCa CI [-0.5, -0.065], p=0.026), postoperative L-Dopa (r = -0.443, 95% BCa CI [-0.624, -0.233], p=0.001), postoperative LEDD (r = -0.404, 95% BCa CI [-0.58, -0.188], p=0.003) (**Figure 2**).

**Figure 2:**
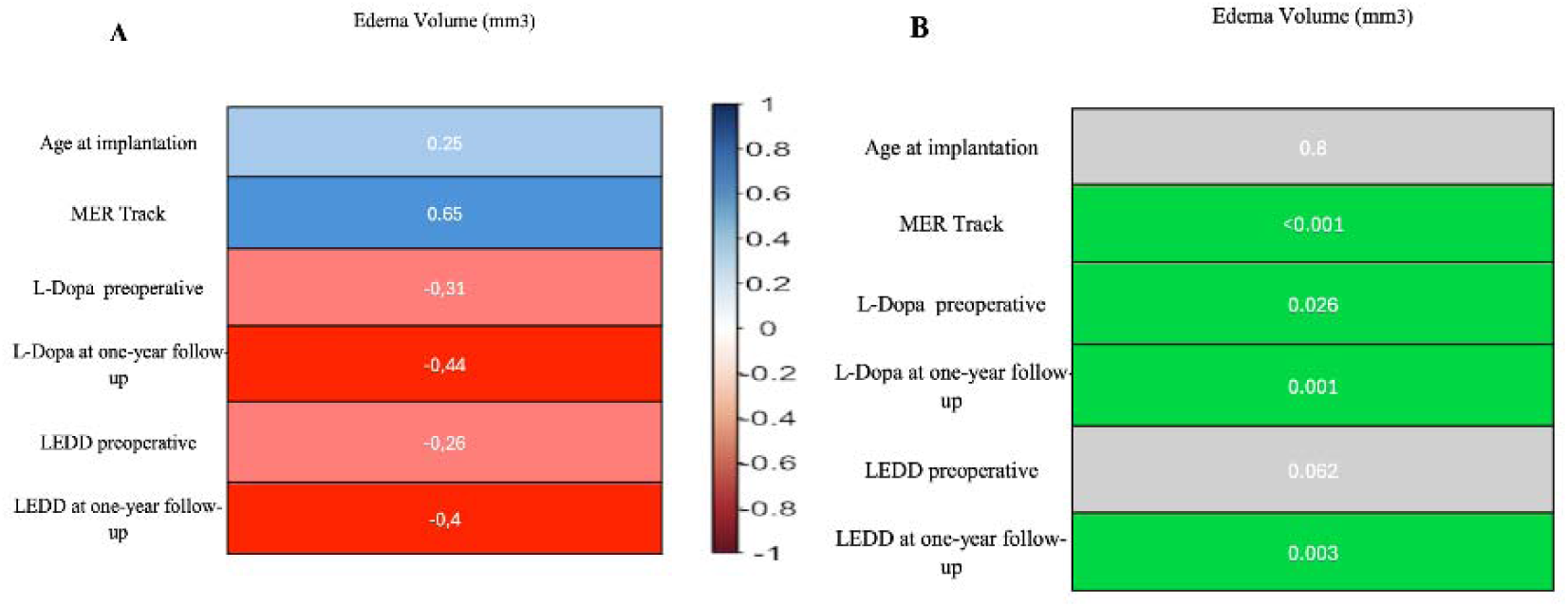
Correlation matrix and significance values from correlation analysis **A:** Correlation matrix showing Pearson’s correlation coefficient (r) from correlation analysis between significant variables and edema volume. On the right, the color-coded scale displays the strength of correlation. **B**: p values from each correlation. Note that only six correlations (enlightened in green) were statistically significant. MER: microelectrode recordings. L-Dopa: levodopa dosage in mg. LEDD: levodopa equivalent daily dose.

However, the number of MER tracks was the only variable that significantly contributed to the model in a multiple regression analysis (adjusted R^2^ = 0.408, F(1,49) = 35.47, p<0.001). Specifically, MER tracks explained 65% of the variance in edema volume (unstandardized B = 10.204 ± 1.713, β = 0.65, p<0.001, sr^2^ = 0.65).

Non-significant results from each regression analysis are reported in **Supplementary Materials**.

## Discussion

We analyzed a retrospective cohort of PD patients displaying PLE after STN-DBS and demonstrated that edema had no impact on long-term clinical and neuropsychological outcomes. Nonetheless, the first post-operative MRI showed an edematous reaction around the electrodes in 68.62% of our patients. This result confirms an emerging trend from the current literature, i.e. PLE is a more frequent complication than originally described^12,23^. Several factors may contribute to the discrepancies observed in different studies. Firstly, the imaging modality used to identify PLE can influence the timely diagnosis of this complication. In fact, the first PLE reports were based on CT scans, which are routinely performed in many centers to verify the correct lead placement in the immediate postoperative period. However, it is now unanimously acknowledged that CT is less sensitive than MRI for PLE detection^24^. Furthermore, not all MRI sequences have the same characteristics and T2-weighted sequences can better delineate edematous reactions^25^. In this context, FLAIR is a T2 sequence acquired with an inversion time modality to negate the cerebrospinal fluid’s contribution to the magnetic signal, thus allowing a more precise characterization of fluid collections and edema^26^. Therefore, we decided to employ the FLAIR sequence to estimate PLE in our patients. Secondly, the timing of postoperative imaging acquisition seems to play a not negligible role in PLE identification. In one of the earliest PLE series, Englot et al.^27^ observed that PLE was present in only 1% of patients at day-1 brain MRI, while PLE reached a 50% incidence if the MRI scan was performed between the 5th and 30th postoperative days. More recently, our group prospectively followed 19 consecutive PD patients after STN-DBS and demonstrated that all subjects had PLE in at least one hemisphere on consecutive MRI between the 7th and 20th postoperative days^14^. Nevertheless, we found no significant differences in postoperative MRI scheduling time between patients with and without PLE. This result underscores that PLE is not a simply radiological epiphenomenon, but it is influenced by patient- and surgery-related factors.

The etiology of PLE remains unclear. While all studies agree on its non-infectious nature after extensive laboratory and imaging workup^28,29^, different theories have been formulated regarding the pathophysiology of PLE. Some authors sustained that PLE has an allergic or immune-mediated origin^30^. From this perspective, PLE would have been caused by a hypersensitivity reaction to some components of DBS electrodes or any of the materials employed during the surgery, i.e. irrigation solutions, firing glues or other dural sealants^31,32^. Furthermore, Servello et al.^33^ even reported more frequent edematous reactions with leads from one vendor than others. However, this immune-mediated theory is not supported by the fact that PLE usually self-resolves without steroid therapy despite the persistence of the presumed offending agent and that many patients manifest unilateral brain edema even after bilateral procedures^27^. On the other hand, our results seem to point to the hypothesis of altered blood-brain barrier permeability^12^. According to this view, PLE would be due to mechanical mico-lesioning of the blood-brain barrier and subsequent transient increase of water filtering into the interstitial space^34,35^. In fact, we found that more MER tracks were used in PLE patients than in subjects without edema. Moreover, total edema volume was directly proportional to the total number of MERs, which also resulted in being the only significant predictor of edema volume after controlling for other clinical variables. Although the notion of a more frequent edematous reaction in patients undergoing MER has already been reported^13,36^, no authors presented statistically significant differences or correlations possibly because of the smaller cohorts in the previous series.

Patients with PLE are usually asymptomatic and no pharmacological treatments are needed for this complication that is often purely radiologic^24,36,37^. However, reports exist regarding PLE patients presenting with mental state deterioration within hours after the implantation or with seizures^31,38–40^ and focal neurological deficits along with MRI evidence of cavitation reaction accompanying PLE^18^. In these cases, symptoms regressed after steroid administration^41^. In our cohort, 26% of PLE patients experienced transient episodes of confusion. Usually beginning between 24 and 48 hours after the implantation, this cognitive alteration consisted of a variable mixture of thought slowdown, space and time disorientation and decreased motor initiative. These episodes were self-resolving and did not need medical therapy. However, this subset of PLE patients had a longer hospital stay for further observation.

Together with edema volume, the patient’s age at the implantation was another significant difference between patients with and without edema. Indeed, PLE patients in our cohort were significantly older than patients without edema, although the patient’s age did not significantly correlate with edema volume. In a similar study, Giordano et al.^13^ also used advanced patient age to support the hypothesis of mechanical damage for PLE pathogenesis, i.e. older brain tissue is more fragile and more prone to enhanced vasogenic and inflammatory responses to surgical insults^42,43^. In this context, multiple brain penetration with MER tracks assumes further importance in the possible PLE etiology. Although the usage of more than one MER is linked to clinically relevant brain haemorrhages^7,44^, our patients only presented small hemorrhagic spots, which were clinically silent and no statistically significant differences in hemorrhagic spots resulted between the two patient groups. These findings may be explained if we consider multiple MERs as a necessary but not sufficient factor for PLE occurrence, since other variables – such as advanced age – may contribute. Finally, we found no differences in long-term motor and neuropsychological outcomes between PLE patients and those without edema in accordance with previous studies^23,45^. In fact, DBS was demonstrated not to alter the long-term cognitive profiles of PD patients compared to not-implanted PD controls^46,47^.

### Limitations

The main limitation of our work is its retrospective nature, which may lead to selection bias. While the monocentric nature of our study grants that all patients were treated and followed homogeneously, our results may not be fully generalizable because of some slight differences in surgical technique existing among centers. Furthermore, we implanted the previous model of omnidirectional DBS leads, since the latest directional leads were only recently introduced in our country. In clinical practice, adopting directional leads reduced the need for multiple MERs for some centers or completely abolished their usage for others^48–50^. By so doing, this technical advance would remove one of the PLE risk factors identified in our study. However, the stringent need for MER in DBS procedures is still a matter of debate^51,52^ and prominent groups still sustain the importance of a precise neurophysiological delineation of the target nucleus by MER mapping before permanent electrode implantation^53,54^. Nonetheless, the relationship between directional leads and PLE deserves further investigation and will be the subject of subsequent studies by our group.

In this study, we presented a retrospective cohort of PD patients displaying PLE after STN-DBS and suffering from transient postoperative confusion. We also described the influence of multiple MER tracks on PLE and postulated an important causative role related to it. Based on our results, we suggest limiting the number of MER tracks in older patients, who are known to be prone to delirium and altered mental status after surgical procedures^55^. In fact, several brain penetrations seem to be related to PLE, which causes postoperative confusion with longer hospitalization.

## Data Availability

All data produced in the present study are available upon reasonable request to the authors

