## Supplementary Table for "Peri-Lead Edema in Deep Brain Stimulation: Long-Term Outcomes and Possible Etiological Correlates"

**Supplementary Table:** Non-significant results from regression analysis

| **Dependent variable** | **Added predictor** | **Adjusted R^2^** | **p-value** |
| --- | --- | --- | --- |
| Edema volume | MER Tracks | 0.40 | <0.001 |
|  | L-Dopa pre | 0.43 | 0.091 |
|  | L-Dopa post, LEDD post | 0.45 | 0.184 |
